# Comparative immunogenicity and relative vaccine efficacy of recombinant and cell culture-based influenza vaccines: Results from a randomized trial of adults in the United States, 2024-2025

**DOI:** 10.64898/2026.10.01.26364413

**Authors:** Kelsey M. Sumner, Emma K. Noble, Lauren B. Grant, Jessica R. Meeker, Vel Murugan, Lora Nordstrom, Stacey L. House, Rachel M. Presti, Emmanuel B. Walter, Olivia L. Williams, Elie A. Saade, David H. Canaday, Katherine V. Williams, Richard K. Zimmerman, F. Liaini Gross, Sara Valencia, Brian Gurbaxani, Nathaniel M. Lewis, Zhu-Nan Li, Vasiliy Mishin, Larisa Gubareva, Bin Zhou, Min Z. Levine, Brendan Flannery, Sascha Ellington, the US Flu VE Network Collaborators

**Affiliations:** Influenza Division, US Centers for Disease Control and Prevention, Atlanta, USA; US Public Health Service, Rockville, MD, USA; Arizona State University, Biodesign Center for Personalized Diagnostics, College of Health Solutions and Health Observatory, Arizona State University, Tempe, AZ; Valleywise Health, Phoenix, AZ; Washington University School of Medicine, Department of Emergency Medicine, St. Louis, MO; Washington University School of Medicine, St. Louis, MO; Duke University School of Medicine, Department of Pediatrics, Durham, NC; Duke University School of Medicine, Duke Human Vaccine Institute, Durham, NC; University Hospitals Cleveland Medical Center, Cleveland, OH; Case Western Reserve University School of Medicine, Cleveland, OH; Louis Stokes Veterans Affairs Medical Center, Cleveland, OH; University of Pittsburgh School of Medicine, Pittsburgh, PA

**Keywords:** Influenza, Vaccine efficacy, Recombinant, Cell culture

## Abstract

**Background:** Recombinant (RIV) and cell culture-based inactivated (ccIIV) influenza vaccines could improve seasonal influenza prevention; however, evidence is limited in how protection may differ among non-egg-based vaccines.

**Methods:** We conducted a randomized trial of RIV versus ccIIV to compare vaccine immunogenicity in adults aged 18 to 64 years during the 2024-25 influenza season (ClinicalTrials.gov, NCT06518577). Active surveillance for influenza-like illness was conducted for six months post-vaccination and cumulative incidence of influenza by vaccine type was compared to estimate relative vaccine efficacy. Participants were tested for influenza by reverse-transcription polymerase chain reaction or rapid antigen tests. Immunogenicity was evaluated by hemagglutination inhibition (HAI), virus microneutralization (MN), and neuraminidase inhibition (NAI) assays in a subset of blood specimens collected pre-vaccination and one-month post-vaccination.

**Results:** Among the 591 participants, 25 cases of influenza A were detected; 2.4% cumulative incidence among RIV recipients versus 6.1% among ccIIV recipients. The probability of influenza was 61% lower with RIV than ccIIV (95% confidence interval: 7 to 84). Among 151 participants, post-vaccination HAI and MN titers vaccine components were higher among RIV recipients while NAI titers were higher among ccIIV recipients.

**Conclusions:** RIV provided better protection than ccIIV against influenza A illness in adults during the U.S. 2024-2025 season.

## INTRODUCTION

Seasonal influenza vaccines are traditionally produced from influenza viruses isolated and propagated in embryonated chicken eggs. Influenza immunization favors boosting of pre-existing antibody over generation of new antibody and therefore could increase response against egg-adapted epitopes.^1–4^ Cell culture-based and recombinant influenza vaccines were developed to improve immune response to circulating influenza viruses without egg-adapted mutations as they utilize viruses that were not propagated in eggs.^5–8^ Prior trials have attempted to estimate the relative vaccine efficacy (rVE) of cell culture-based and recombinant influenza vaccines, however these studies observed low attack rates within their immunogenicity subset or had limited statistical power to compare cell culture-based and recombinant vaccines to one another.^8,9^ The two non-egg-based vaccines differ in influenza virus hemagglutinin (HA) protein content and only the cell culture-based influenza vaccine contains neuraminidase (NA) protein.^10,11^ Prior comparative immunogenicity trials have observed a higher HA antibody response among those that receive recombinant influenza vaccine; however, it remains unclear if the higher antibody response observed translates to higher protection against influenza illness.^8,9,12^ We conducted an open-label trial of two trivalent U.S.-licensed influenza vaccines among adults aged 18-64 years randomized to receive either a recombinant influenza vaccine (RIV; Flublok®, Protein Sciences Corporation/Sanofi) or cell culture-based inactivated influenza vaccine (ccIIV; Flucelvax®, Seqiris Inc.) to compare immunogenicity with post-hoc relative efficacy of non-egg-based influenza vaccines during the 2024-2025 influenza season in the United States.

## METHODS

### Trial design and oversight

During the 2024-2025 influenza season, we conducted an open label, randomized comparative immunogenicity trial in an enrolled cohort of adults aged 18-64 years across six study sites in four US states: Arizona, Missouri, Ohio, and Pennsylvania. The study period from first enrollment through end of follow-up was September 3, 2024 to June 4, 2025.

The protocol was reviewed and approved by the Duke University Health System Institutional Review Board with participating study sites and the US Centers for Disease Control and Prevention (CDC) under reliance arrangements (See 45 C.F.R. part 46.114; 21 C.F.R. part 56.114). The trial protocol was made publicly available on ClinicalTrials.gov (NCT06518577) on July 24, 2024.^13^ All participants provided written informed consent.

CDC investigators had primary responsibility for trial design, protocol development, immunogenicity and NA content testing, and data analysis; the coordinating center (Duke University) for trial monitoring and data management; and the investigators at the trial sites for critical protocol review, trial procedures, and data collection. All authors assume responsibility for the accuracy and completeness of the data and for the fidelity of the trial to the protocol. Consolidated Standard of Reporting Trials (CONSORT) reporting guidelines were followed in preparation of this publication.^14^

### Trial population and randomization

Adults aged 18-64 years who had not received a 2024-2025 Northern Hemisphere influenza vaccine were randomized (1:1) to receive either RIV or ccIIV at enrollment (day 1). Exclusion criteria included history of a severe allergic reaction after a previous dose of any influenza vaccine or vaccine component, receipt of any live vaccine within 6 weeks or non-live vaccine within 2 weeks prior to enrollment, history of Guillain-Barré syndrome, pregnancy, or bleeding diathesis. A full list of inclusion and exclusion criteria can be found in the publicly available protocol.^13^ Randomization group was assigned to patients using computer-generated permuted block randomization stratified by enrollment site. Participants, trial staff, and investigators were unblinded to randomization assignments.

### Vaccines

RIV contains 45 μg of recombinant HA protein per influenza strain in a 0.5-mL dose (i.e., a total of 135 µg of HA in a trivalent dose), produced using a baculovirus expression vector system in insect cells, and no NA protein. ccIIV contains 15 µg of HA per influenza strain in a 0.5-mL dose (i.e., a total of 45 µg of HA in a trivalent dose) with the viruses propagated in Madin-Darby canine kidney cells.^11^ ccIIV also contains NA protein, however NA content is not regulated and can vary by influenza subtype and vaccine lot.^15–17^ Both vaccines contained each of the two influenza type A subtypes (A(H1N1)pdm09 and A(H3N2)) and influenza type B Victoria lineage, as recommended by FDA’s Vaccines and Related Biological Products Advisory Committee for the 2024-2025 season.^10,11,18^ RIV was approved and recommended for persons aged ≥18 years and ccIIV for persons aged ≥6 months.^10,11,19^ Vaccine was supplied in pre-filled syringes. After randomization, vaccines were administered at the enrollment visit intramuscularly by licensed healthcare personnel. NA content quantification was conducted on the ccIIV lot distributed to sites using mass spectrophotometry as previously published;^20^ all participants received the same vaccine lot of ccIIV to standardize NA protein content across sites. RIV vaccine does not contain NA and therefore NA was not quantified.

### Immunogenicity

Blood samples were drawn at enrollment (before vaccination), and approximately 28 (28-42) days and 180 (150-210) days after vaccination to measure immune responses to vaccination. The main objective of the trial was to determine and compare pre– and post-vaccination influenza antibody titers by HAI for both vaccination groups (in a random subset of participants). Participants with both pre– and 28 days post-vaccination serum results and no documented influenza illness in between these visits were eligible for the subset. The immunogenicity subset was selected using simple random sampling by site and vaccine randomization group and powered based on 80% power to detect a difference in seroconversion levels assuming 41% seroconversion for RIV recipients and 21% seroconversion for ccIIV recipients.^8,12^ The subset of collected sera were tested for influenza HA protein antibody titers against cell culture-propagated 2024-2025 Northern hemisphere vaccine reference strains: A/Wisconsin/67/2022 [A(H1N1)pdm09-like], A/Massachusetts/18/2022 [A(H3N2)-like], and B/Austria/1359417/2021 [B/Victoria lineage-like]. Sera were tested by hemagglutination inhibition (HAI) for A(H1N1)pdm09 and B/Victoria strains and by microneutralization (MN) for A(H3N2) strain using validated protocols.^21,22^ MN assay was used in lieu of HAI for the A(H3N2) strain as this assay is known to be more sensitive, particularly when testing against cell-grown A(H3N2) strains.^23^ Influenza B virus antigen was ether-treated for HAI assays. Sera were also tested for neuraminidase inhibition (NAI) titers using the enzyme-linked lectin assay against recombinant A(H6) viruses, A/Wisconsin/67/2022 [A(H6N1)] and A/Massachusetts/18/2022 [A(H6N2)], to minimize interference of HA-specific antibodies.^24,25^ NAI titers were not measured for B/Victoria lineage.

### Influenza relative vaccine efficacy

Participants responded to a weekly automatic electronic survey regarding the occurrence of a new onset of influenza-like illness (ILI), defined as onset in the past 7 days of fever and cough or sore throat (lasting ≥24 hours), and any change in health status requiring hospitalization. Influenza test results were also reported if available and may have been from at-home tests or testing in a healthcare facility unrelated to study procedures. Participants who reported ILI were asked to provide a respiratory specimen for standard point-of-care respiratory virus testing (reverse transcription-polymerase chain reaction [RT-PCR] or rapid antigen test) within 10 days of symptom onset. In addition, up to two blood draws were collected (within 10 days of symptom onset and 28 days after a positive influenza test). Influenza A infection was determined by RT-PCR or rapid antigen testing; subtyping was determined by either RT-PCR or serologic response to infection (**Supplemental Methods**).

### Safety

The trial evaluated two licensed influenza vaccines with previously described reactogenicity and safety profiles.^10,11,26,27^ Only serious adverse events (SAEs) that occurred during the study period were recorded (**Supplemental Methods**). SAEs were reported to study staff by participants and site principal investigators determined relationship to study vaccine or procedures. Data on reactogenicity and non-serious adverse events were not obtained.

### Statistical analysis

For each vaccine antigen, we compared the geometric mean titer (GMT) of sera collected pre– and 28 days post-vaccination in antibody produced by HAI, MN, or NAI testing within each randomization group as well as geometric mean fold rise (GMFR). A linear regression model adjusted for study site compared pre– and post-vaccination GMTs and GMFR by randomization group. Percent seroconversion and seroprotection for HAI and MN titers were also compared by randomization group (**Supplemental Methods**). Titers were log_2_-transformed prior to analysis for all assays.

A post-hoc analysis estimated rVE of RIV compared to ccIIV against RT-PCR or rapid antigen test confirmed influenza illness. We included participants who did not have any influenza illness within 14 days of vaccination and completed at least one weekly survey 14 or more days after vaccination (efficacy population). Post-hoc power calculations estimated that the data on 591 participants eligible for efficacy analysis had 65% power to demonstrate relative effectiveness of RIV compared to ccIIV with a 2-sided alpha=0.05, rVE of 60% and influenza rate of 6.4% among those receiving ccIIV. We used Cox proportional hazards models to estimate hazard ratios (HR) and calculated rVE as (1 – HR), expressed as a percentage. The proportional hazards assumption for the Cox proportional hazards models was assessed using Kaplan-Meier curves and Schoenfeld residual plots. Sensitivity analyses considered alternative endpoints for influenza illness (e.g., self-reported illness or RT-PCR-positive infections) as well as a model including enrollment site as a stratum.

Analyses were performed in R version 4.3.1 software (R Core Team, Vienna, Austria). Statistical significance was assessed at an α-level of 0.05.

## RESULTS

### Analytic populations

From September 3 to November 11, 2024, 606 participants enrolled in the trial, 599 were randomized to receive RIV or ccIIV, and 595 received their randomization vaccine (**Figure 1**). A total of 151 participants were randomly selected for the immunogenicity subset (76 received RIV and 75 received ccIIV) and 591 participants were in the efficacy population (294 received RIV and 297 received ccIIV). Baseline characteristics and demographics were similar between randomization groups in both populations (**Table 1 and Supplemental Table 1**). Three SAEs were identified during follow-up but were all determined to be unrelated to study vaccine or procedures (**Supplemental Table 4**).

**Figure 1.**
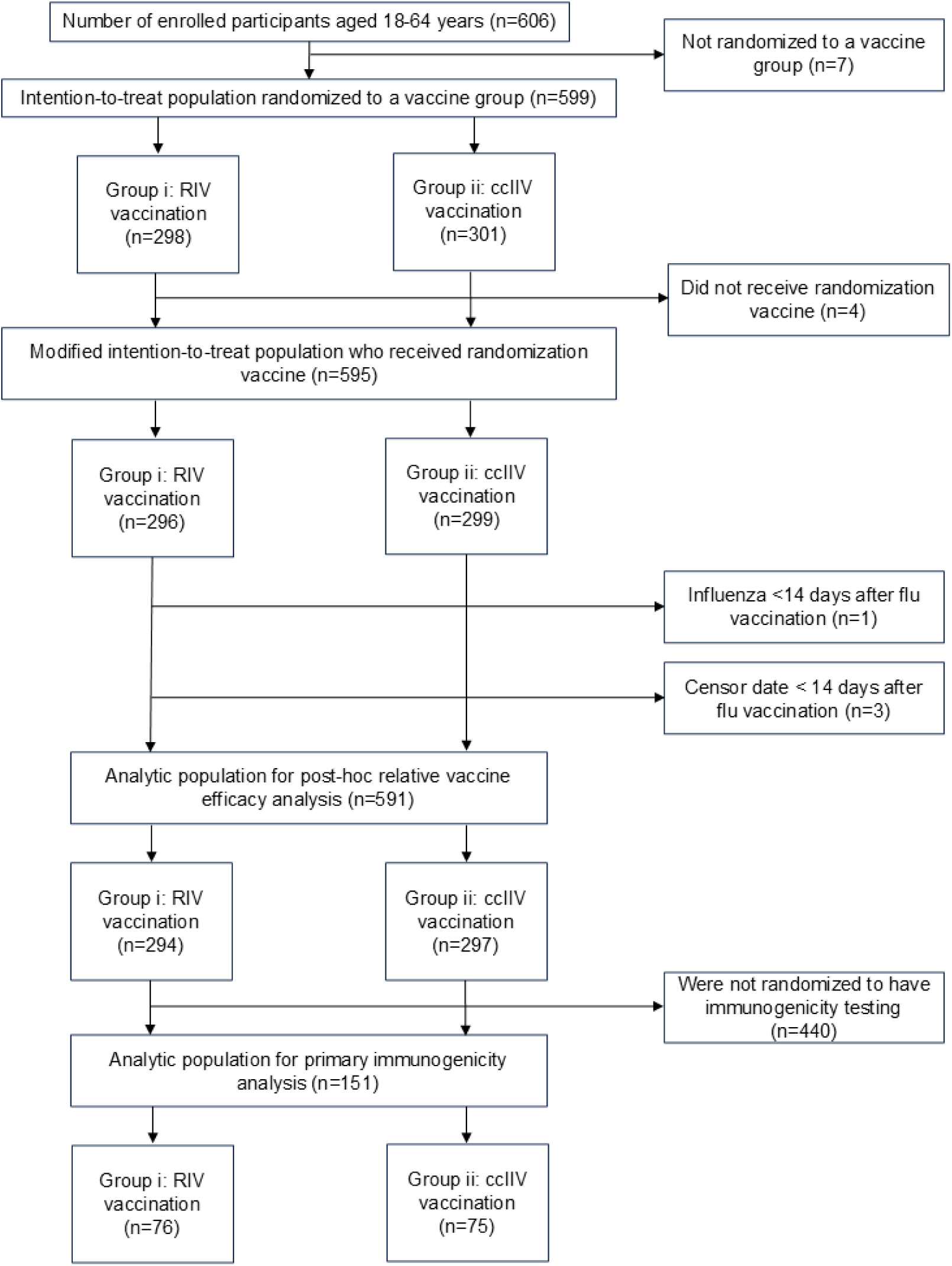
Total number of enrolled participants by randomization group. Participants were randomized to receive one of two influenza vaccines: ccIIV or RIV. Abbreviations: ccIIV, cell culture-based inactivated influenza vaccine; RIV, recombinant influenza vaccine

**Table 1.** Demographics of study participants in the relative vaccine efficacy analytic population.

| Characteristic | Randomization group |  |
| --- | --- | --- |
|  | RIV | cclIV |
| **Total N** | 294 | 297 |
| **Age (years), median (IQR)** | 40 (28, 52) | 37 (28, 47) |
| **Age Group, n (%)** |  |  |
| 18-49 years | 208 (70.7%) | 231 (77.8%) |
| 50-64 years | 86 (29.3%) | 66 (22.2%) |
| **Sex,<sup>a</sup> n (%)** |  |  |
| Female | 203 (69.0%) | 202 (68.0%) |
| Male | 88 (29.9%) | 95 (32.0%) |
| **Race/ethnicity, n (%)** |  |  |
| Hispanic or Latino | 33 (11.2%) | 40 (13.5%) |
| American Indian or Alaska Native | 0 (0.0%) | 2 (0.7%) |
| Asian | 37 (12.6%) | 41 (13.8%) |
| Black or African American | 17 (5.8%) | 17 (5.7%) |
| Middle Eastern or North African | 0 (0.0%) | 3 (1.0%) |
| Multiracial | 9 (3.1%) | 5 (1.7%) |
| Native Hawaiian or Other Pacific Islander | 0 (0.0%) | 0 (0.0%) |
| White | 194 (66.0%) | 187 (63.0%) |
| Other | 2 (0.7%) | 1 (0.3%) |
| Unknown | 2 (0.7%) | 1 (0.3%) |
| **Enrollment Site, n (%)** |  |  |
| Arizona State University | 66 (22.4%) | 68 (22.9%) |
| Valleywise Health Medical Center | 22 (7.5%) | 22 (7.4%) |
| Washington University | 75 (25.5%) | 74 (24.9%) |
| University Hospitals | 69 (23.5%) | 71 (23.9%) |
| Cleveland Veterans Affairs | 13 (4.4%) | 12 (4.0%) |
| University of Pittsburgh | 49 (16.7%) | 50 (16.8%) |
| **Prior season influenza vaccination, n (%)** |  |  |
| No | 27 (9.2%) | 23 (7.7%) |
| Yes | 259 (88.1%) | 270 (90.9%) |
| I don't know | 8 (2.7%) | 4 (1.3%) |
| **Current season influenza A illness,<sup>b</sup> n (%)** |  |  |
| Negative | 287 (97.6%) | 279 (93.9%) |
| Positive<sup>c</sup> | 7 (2.4%) | 18 (6.1%) |
| **Included in immunogenicity subset,<sup>d</sup> n (%)** | 76 (25.9%) | 75 (25.3%) |

Abbreviations: cclIV, cell culture-based inactivated influenza vaccine; RIV, recombinant influenza vaccine
<sup>a</sup> Sex was unknown for 3 participants; 3 RIV
<sup>b</sup> Defined as a test-positive influenza A illness as indicated by a positive molecular or rapid diagnostic test result.
<sup>c</sup> Influenza confirmed by at-home antigen test for 4 participants; 2 RIV and 2 cclIV
<sup>d</sup> The distribution of participants randomized to receive RIV versus cclIV across demographics for the immunogenicity subset (N=151) is found in **Supplemental table 1**.

### Immunogenicity

Across vaccine components, no difference was observed in pre-vaccine HAI, MN, or NAI GMTs by randomization group (**Table 2**), except for pre-vaccine NAI titers against influenza A(H1N1)pdm09, which were higher among participants receiving ccIIV compared to those receiving RIV. Post-vaccine hemagglutinin response (HAI or MN titers) against all vaccine components was higher in RIV compared to ccIIV recipients (**Table 2**). After quantification of NA content in the vaccine lot given to those randomized to receive ccIIV, we found there to be <0.5 µg for A(H1N1)pdm09, 10.4 µg for A(H3N2), and 1.5 µg for influenza B per dose of ccIIV. Post-vaccine neuraminidase response against influenza A(H1N1)pdm09 and A(H3N2) was higher for ccIIV recipients (**Table 2**).

**Table 2.**
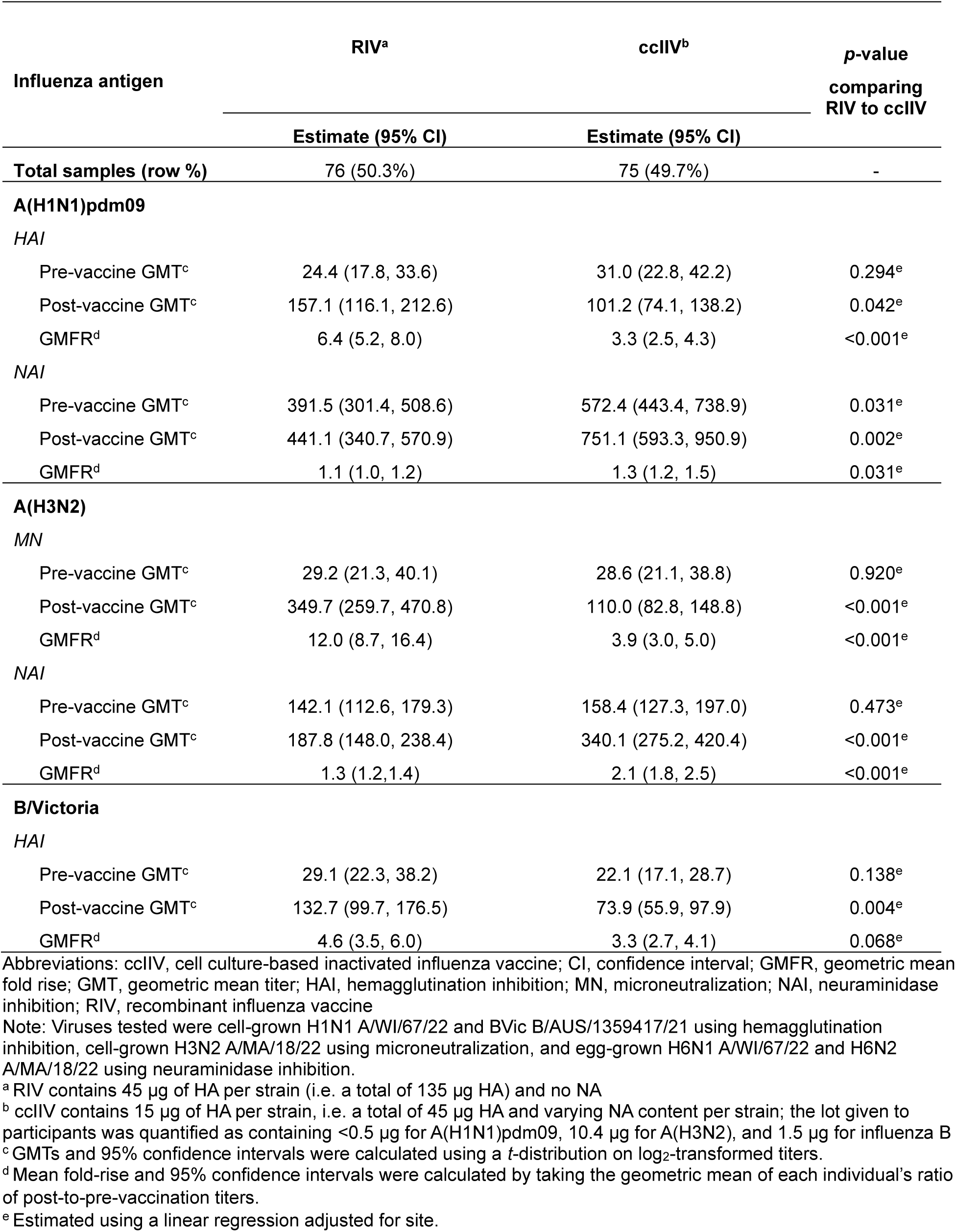
Comparison of RIV and ccIIV influenza vaccine response using hemagglutination inhibition, microneutralization, or neuraminidase inhibition titers.

### Influenza relative vaccine efficacy

Participants were followed for detection of influenza illness for a median of 168 days (range: 12 to 238 days). Overall adherence to weekly surveys was 95% in both vaccine groups. Among participants reporting one or more ILI episodes, 43% completed study swabs and 62% either completed a study swab or self-reported an influenza test. Twenty-five participants tested positive for influenza A, including 8 A(H1N1)pdm09, 11 A(H3N2), 1 co-infection with A(H1N1)pdm09 and A(H3N2), and 5 not subtyped. No influenza B cases were detected although 1 was self-reported via the weekly surveys. The cumulative incidence of influenza A illness was 2.4% (7/294) among RIV recipients and 6.1% (18/297) among ccIIV recipients (**Figure 2)**.

**Figure 2.**
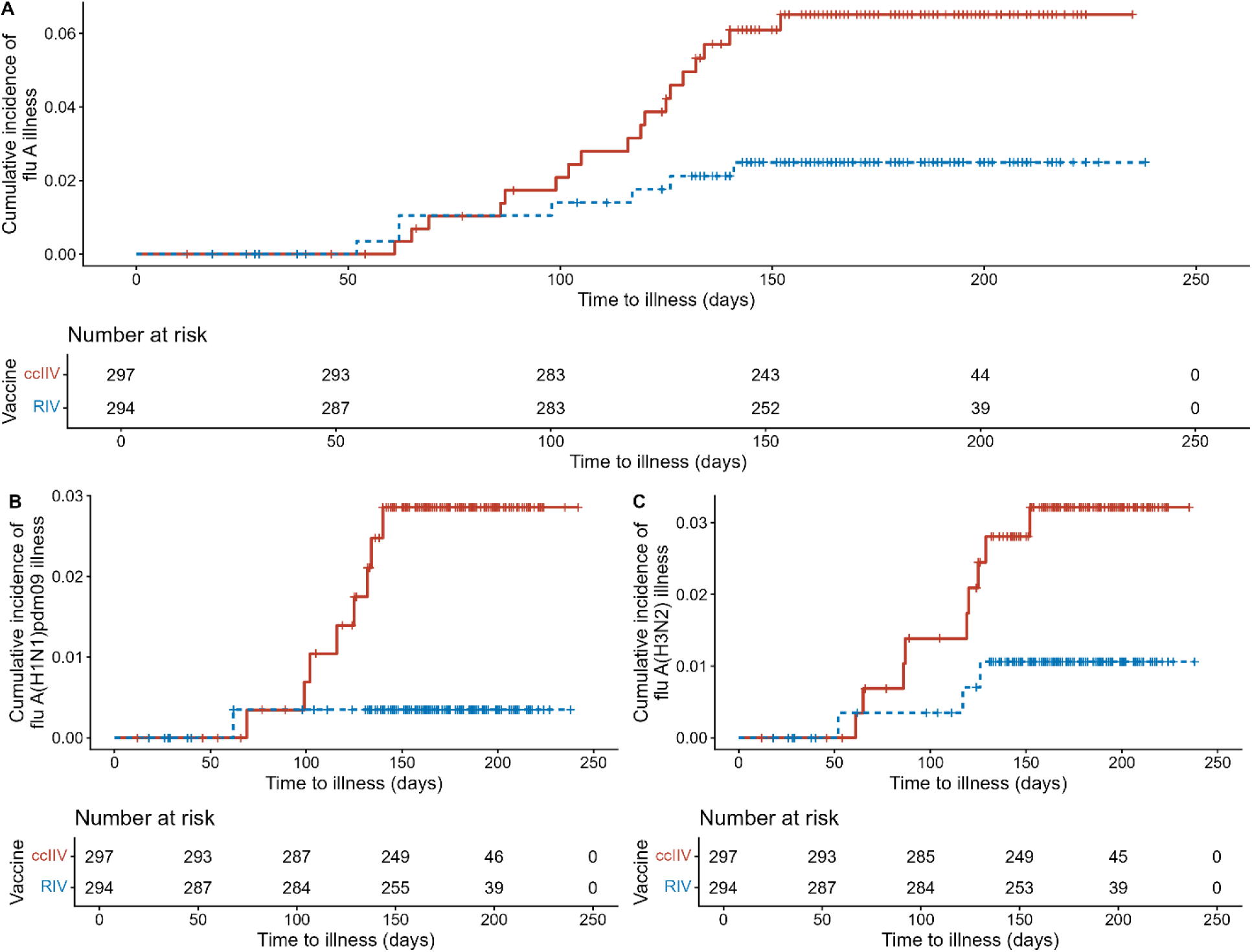
Cumulative incidence of influenza A overall (panel A), influenza A(H1N1)pdm09 (panel B), and influenza A(H3N2) (panel C) from the time to illness onset (in days). Cumulative incidence curves are presented stratified by influenza vaccine received (ccIIV in red or RIV in blue) with number at risk at 50-day intervals shown below. Abbreviations: ccIIV, cell culture-based inactivated influenza vaccine; RIV, recombinant influenza vaccine

The rVE of RIV compared with ccIIV against any confirmed influenza A illness was 61% (95% CI: 7 to 84%). rVE was 87% (95% CI: –1 to 98%) against A(H1N1)pdm09 and 66% (95% CI: –24 to 91%) against influenza A(H3N2) (**Figure 3**). Relative efficacy was similar in sensitivity analyses of alternative endpoints and model adjustments (**Supplemental Table 3**).

**Figure 3.**
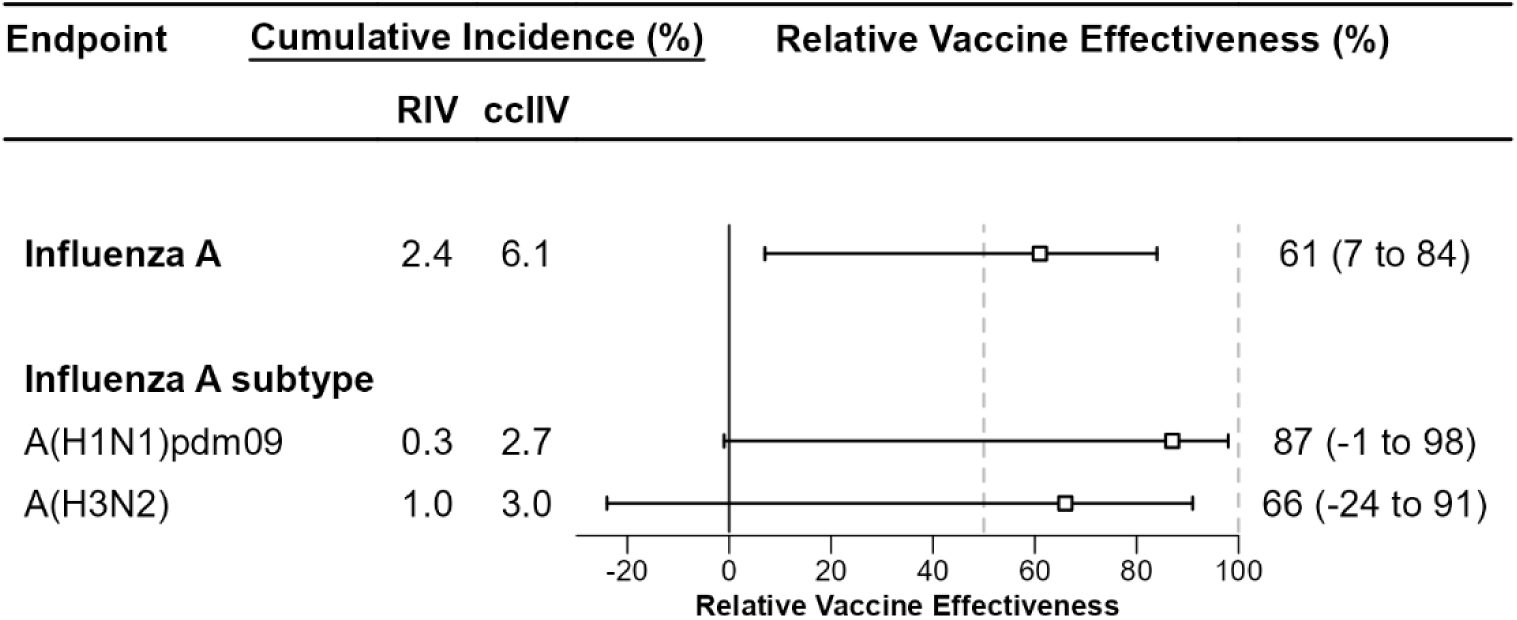
Relative vaccine efficacy against influenza A overall, influenza A(H1N1)pdm09, and influenza A(H3N2) comparing receipt of RIV to ccIIV. Relative vaccine efficacy (rVE) of RIV compared to ccIIV was calculated as 1 minus the hazard ratio produced by running a Cox proportional hazards model of the time to influenza illness adjusted for influenza vaccine type. The rVE was multiplied by 100 to estimate percentages. Note that subtype results were not available for 6 of 26 influenza-positive cases; these 6 participants were censored at the time of symptom onset for the influenza A subtype-specific rVE estimates. Abbreviations: ccIIV, cell culture-based inactivated influenza vaccine; CI, confidence interval; RIV, recombinant influenza vaccine

## DISCUSSION

In this randomized comparative immunogenicity trial among U.S. adults aged 18 to 64 years, RIV elicited higher influenza virus neutralizing or hemagglutination inhibiting antibodies and was associated with higher vaccine efficacy against symptomatic influenza A virus infection compared with ccIIV. Post-vaccine neuraminidase antibody response was higher for ccIIV recipients. The 2024-2025 influenza season was classified as a high severity season exhibiting high incidence of influenza illness in the United States^28,29^ and co-circulation of influenza A subtypes A(H1N1)pdm09 and A(H3N2),^30^ with lower rates among those receiving RIV than ccIIV in this trial. RIV provided 61% higher protection against influenza A illness compared with ccIIV. rVE estimates by influenza A subtype were similar to the overall flu A estimate but not statistically significant, possibly due to small sample size. The greater protection conferred by RIV might be because it contains three times more HA protein per strain than ccIIV.

This immunogenicity study demonstrated both higher antibody response against cell-propagated influenza virus antigens and associated differences in observed protection against confirmed influenza A between two non-egg-based influenza vaccines. Higher antibody responses following RIV have been observed in prior immunogenicity studies when comparing to only egg-based influenza vaccines^8,31^ or both cell culture– and egg-based influenza vaccines.^7,12^ Only one trial measured NA response in RIV, standard dose, high-dose, and adjuvanted influenza vaccines and found that NA response was higher in other influenza vaccines than RIV,^32^ as expected due to the lack of NA protein in RIV. Other trials and observational studies have observed an rVE of 15–30% among recipients of RIV, however these studies only compared RIV to standard-dose egg-based vaccines.^8,9,31^ One trial found no significant rVE between ccIIV or RIV when compared with standard-dose egg-based vaccines. However it did not compare ccIIV and RIV to each other, and enrollment overlapped with the COVID-19 pandemic which may have influenced estimates due to low influenza circulation.^33^ In addition to our rVE estimates utilizing different comparator vaccines, rVE estimates are sensitive to absolute vaccine efficacy^34^ and therefore the season in which the trials are conducted may have contributed to differences in observed rVE.

Results suggest that purified HA protein (in a dose of 135 µg) produced in insect cells elicited virus neutralizing and hemagglutination inhibition antibodies associated with protection against influenza illness. RIV, which contains no neuraminidase protein, provided a control for protection from only pre-existing levels of neuraminidase-inhibiting antibody rather than vaccine-induced NAI antibody response, although it does not address whether including NA protein in RIV or increasing NA content in ccIIV would have improved protection. However, the point estimate for rVE was lower against influenza A(H3N2) than against A(H1N1)pdm09, suggesting there may be some additional protection offered by NA in the vaccine as there was detectable levels of NA2 protein but not NA1 in the lot of ccIIV received by participants.^15–17^ Immunity against NA has been identified as an important component of protection against influenza.^35,36^ Previous immunogenicity studies comparing RIV with ccIIV have not measured NAI antibodies. In this study, pre-existing NA antibodies or cell-mediated immune responses from previous influenza virus infection or vaccination may have contributed to protection. Quantification of virus neuraminidase content in ccIIV was consistent with two-fold increase in NA2-specific neuraminidase inhibition assay titers and low response against NA1-specific antibodies following vaccination, based on undetectable NA1 content in the ccIIV lot participants received.

This trial had several limitations. First, respiratory virus testing was not performed for all episodes of respiratory illness reported during follow-up, and respiratory specimens were not systematically collected to detect asymptomatic influenza virus infections. Some infections may have been missed; however, we believe this would have biased rVE estimates towards the null as swab adherence from reported ILI did not differ by vaccination group. Second, participants at one site who reported ILI were tested using rapid antigen detection assays, which may have led to false positive or false negative results. A sensitivity analysis restricting to only sites with RT-PCR testing found similar results to the main analysis. Third, influenza A subtyping was not available for all influenza A-positive specimens, and these unsubtyped participants were censored from analyses at the time of symptom onset for subtype-specific estimates. Fourth, sample size was limited for estimating rVE, particularly against influenza A(H1N1)pdm09 and A(H3N2). Fifth, immunogenicity outcomes were limited to serologic assays; future studies could assess cell-mediated immunity. Finally, results are not generalizable to younger or older age groups and may not be generalizable to other influenza seasons as the study was only conducted in a single season.

To date, this is the first trial to estimate rVE against symptomatic influenza illness between two non-egg-based vaccines in adults aged 18 to 64 years and pair it with immunogenicity measurements of both HA and NA response post-vaccination. Immunity against NA has been identified as an important component of protection against influenza illness;^35,36^ however, our results suggest robust vaccine-induced HA antibody responses without NA antibody response could provide protection against influenza illness as RIV, which contains three times the HA content as ccIIV and no NA content, had higher vaccine efficacy against influenza A illness compared to ccIIV. It is also possible that pre-existing NA antibodies or cell-mediated immune responses may play an additive role in protection. Future work could further investigate the role of humoral and cell-mediated immune response in the greater protection conferred by recombinant influenza vaccines, particularly as new technologies such as mRNA influenza vaccines are being developed.^37^

## ACKNOWLEDGMENTS

Thank you to study participants.

## AUTHOR CONTRIBUTIONS

KMS, EKN, LBG, NL, BF, and SE conceptualized the manuscript. KMS, EN, and LBG conducted statistical analyses. KMS, LBG, EKN, BG, and NL provided methodology. KMS, EKN, VM, LN, SH, RMP, EBW, OLW, EAS, DHC, KVW, RKZ, and BF provided study design, investigation, and coordination. BF and SE provided supervision. FLG, SV, ZL, VM, LG, BZ, and MZL tested specimens. KMS wrote the initial version of the manuscript. All authors read and approved the final version of the manuscript.

## DISCLAIMER

The findings and conclusions in this report are those of the authors and do not necessarily represent the official position of the U.S. Centers for Disease Control and Prevention (CDC). Vaccine trade names and company names are included for informational purposes only and do not represent an endorsement by the CDC and Health and Human Services (HHS).

Dr. Sumner is currently employed by GlaxoSmithKline; however, all work occurred while she was an employee of CDC. Dr. Lewis is currently employed by Merck; however, all work occurred while he was an employee of CDC.

## DATA AVAILABILITY

All data without personal identifiers produced in the present study are available at reasonable request to the corresponding author.

## References

1. Zost SJ, Parkhouse K, Gumina ME, et al. Contemporary H3N2 influenza viruses have a glycosylation site that alters binding of antibodies elicited by egg-adapted vaccine strains. Proc Natl Acad Sci U S A 2017;114(47):12578–12583. DOI: 10.1073/pnas.1712377114.

2. Gambaryan AS, Robertson JS, Matrosovich MN. Effects of Egg-Adaptation on the Receptor-Binding Properties of Human Influenza A and B Viruses. Virology 1999;258:8.

3. Xu X, Kilbourne ED, Hall HE, Cox NJ. Nonimmunoselected Intrastrain Genetic Variation Detected in Pairs of High-Yielding Influenza A(H3N2) Vaccine and Parental Viruses. The Journal of Infectious Diseases 1994;170:7.

4. Raymond DD, Stewart SM, Lee J, et al. Influenza immunization elicits antibodies specific for an egg-adapted vaccine strain. Nat Med 2016;22(12):1465–1469. DOI: 10.1038/nm.4223.

5. Richards KA, Moritzky S, Shannon I, et al. Recombinant HA-based vaccine outperforms split and subunit vaccines in elicitation of influenza-specific CD4 T cells and CD4 T cell-dependent antibody responses in humans. NPJ Vaccines 2020;5:77. DOI: 10.1038/s41541-020-00227-x.

6. Yu ED, Grifoni A, Sutherland A, et al. Balanced Cellular and Humoral Immune Responses Targeting Multiple Antigens in Adults Receiving a Quadrivalent Inactivated Influenza Vaccine. Vaccines (Basel) 2021;9(5). DOI: 10.3390/vaccines9050426.

7. Gaglani M, Kim SS, Naleway AL, et al. Effect of Repeat Vaccination on Immunogenicity of Quadrivalent Cell-Culture and Recombinant Influenza Vaccines Among Healthcare Personnel Aged 18-64 Years: A Randomized, Open-Label Trial. Clin Infect Dis 2023;76(3):e1168–e1176. DOI: 10.1093/cid/ciac683.

8. Grant L, Whitaker JA, Yoon SK, et al. Relative Effectiveness and Immunogenicity of Quadrivalent Recombinant Influenza Vaccine Versus Egg-Based Inactivated Influenza Vaccine Among Adults Aged 18-64 Years: Results and Experience From a Randomized, Double-Blind Trial. Open Forum Infect Dis 2024;11(10):ofae559. DOI: 10.1093/ofid/ofae559.

9. Dunkle LM, Izikson R, Patriarca P, et al. Efficacy of Recombinant Influenza Vaccine in Adults 50 Years of Age or Older. N Engl J Med 2017;376(25):2427–2436. DOI: 10.1056/NEJMoa1608862.

10. US Food and Drug Administration. Package Insert – Flublok. July 01, 2024. (https://web.archive.org/web/20240831173250/, https://www.fda.gov/media/179778/download?attachment).

11. US Food and Drug Administration. Package Insert – Flucelvax. July 01, 2024. (https://web.archive.org/web/20240816041703/, https://www.fda.gov/media/85322/download).

12. Dawood FS, Naleway AL, Flannery B, et al. Comparison of the Immunogenicity of Cell Culture-Based and Recombinant Quadrivalent Influenza Vaccines to Conventional Egg-Based Quadrivalent Influenza Vaccines Among Healthcare Personnel Aged 18-64 Years: A Randomized Open-Label Trial. Clin Infect Dis 2021;73(11):1973–1981. DOI: 10.1093/cid/ciab566.

13. Duke University. Measuring Immunity Against Circulating Influenza Viruses: Randomized Immunogenicity Study Among US Adults Aged 18-64 Years Comparing Two Approved Influenza Vaccines. In: Centers for Disease C, Prevention, Arizona State U, et al., eds. 2024.

14. Hopewell S, Chan AW, Collins GS, et al. CONSORT 2025 Statement: Updated Guideline for Reporting Randomized Trials. JAMA 2025;333(22):1998–2005. DOI: 10.1001/jama.2025.4347.

15. Kang H, Gao J, Giurgea L, Memoli MJ, Daniels R. ELISA-based quantification of neuraminidase in commercial influenza vaccines using virus-derived reference antigens. Microbiology Spectrum 2026. DOI: 10.1128/spectrum.01227-26.

16. Krammer F, Fouchier RAM, Eichelberger MC, et al. NAction! How Can Neuraminidase-Based Immunity Contribute to Better Influenza Virus Vaccines? mBio 2018;9(2):e02332–17. DOI: 10.1128/mbio.02332-17.

17. US Food and Drug Administration. Influenza neuraminidase antigenicity and efficacy in vaccines. (https://www.fda.gov/vaccines-blood-biologics/science-research-biologics/influenza-neuraminidase-antigenicity-and-efficacy-vaccines).

18. US Food and Drug Administration. Use of Trivalent Influenza Vaccines for the 2024-2025 U.S. Influenza Season. (https://www.fda.gov/vaccines-blood-biologics/lot-release/use-trivalent-influenza-vaccines-2024-2025-us-influenza-season).

19. Grohskopf LA, Ferdinands JM, Blanton LH, Broder KR, Loehr J. Prevention and Control of Seasonal Influenza with Vaccines: Recommendations of the Advisory Committee on Immunization Practices – United States, 2024-25 Influenza Season. MMWR Recomm Rep 2024;73(5):1–25. DOI: 10.15585/mmwr.rr7305a1.

20. Williams TL, Luna L, Guo Z, et al. Quantification of influenza virus hemagglutinins in complex mixtures using isotope dilution tandem mass spectrometry. Vaccine 2008;26(20):2510–20. DOI: 10.1016/j.vaccine.2008.03.014.

21. World Health Organization. Manual for the laboratory diagnosis and virological surveillance of influenza. 2011. (https://www.who.int/publications-detail-redirect/manual-for-the-laboratory-diagnosis-and-virological-surveillance-of-influenza).

22. Gross FL, Bai Y, Jefferson S, Holiday C, Levine MZ. Measuring Influenza Neutralizing Antibody Responses to A(H3N2) Viruses in Human Sera by Microneutralization Assays Using MDCK-SIAT1 Cells. J Vis Exp 2017(129). DOI: 10.3791/56448.

23. Heeringa M, Leav B, Smolenov I, Palladino G, Isakov L, Matassa V. Comparability of Titers of Antibodies against Seasonal Influenza Virus Strains as Determined by Hemagglutination Inhibition and Microneutralization Assays. Journal of Clinical Microbiology 2020;58(9):10.1128/jcm.00750-20. DOI: doi:10.1128/jcm.00750-20.

24. Couzens L, Gao J, Westgeest K, et al. An optimized enzyme-linked lectin assay to measure influenza A virus neuraminidase inhibition antibody titers in human sera. J Virol Methods 2014;210:7–14. DOI: 10.1016/j.jviromet.2014.09.003.

25. Westgeest KB, Bestebroer TM, Spronken MI, et al. Optimization of an enzyme-linked lectin assay suitable for rapid antigenic characterization of the neuraminidase of human influenza A(H3N2) viruses. J Virol Methods 2015;217:55–63. DOI: 10.1016/j.jviromet.2015.02.014.

26. Jordan K, Murchu EO, Comber L, et al. Systematic review of the efficacy, effectiveness and safety of cell-based seasonal influenza vaccines for the prevention of laboratory-confirmed influenza in individuals ≥18 years of age. Reviews in Medical Virology 2023;33(3). DOI: 10.1002/rmv.2332.

27. O Murchu E, Comber L, Jordan K, et al. Systematic review of the efficacy, effectiveness and safety of recombinant haemagglutinin seasonal influenza vaccines for the prevention of laboratory-confirmed influenza in individuals ≥18 years of age. Reviews in Medical Virology 2023;33(3). DOI: 10.1002/rmv.2331.

28. Biggerstaff M, Kniss K, Jernigan DB, et al. Systematic Assessment of Multiple Routine and Near Real-Time Indicators to Classify the Severity of Influenza Seasons and Pandemics in the United States, 2003-2004 Through 2015-2016. Am J Epidemiol 2018;187(5):1040–1050. DOI: 10.1093/aje/kwx334.

29. Centers for Disease Control and Prevention. Past Flu Season Severity Assessments. (https://www.cdc.gov/flu-burden/php/surveillance/past-seasons.html).

30. Centers for Disease Control and Prevention. Influenza Activity in the United States during the 2024–25 Season and Composition of the 2025–26 Influenza Vaccine. 2025 (https://www.cdc.gov/flu/whats-new/2025-2026-influenza-activity.html).

31. Dunkle LM, Izikson R, Patriarca PA, Goldenthal KL, Muse D, Cox MMJ. Randomized Comparison of Immunogenicity and Safety of Quadrivalent Recombinant Versus Inactivated Influenza Vaccine in Healthy Adults 18-49 Years of Age. J Infect Dis 2017;216(10):1219–1226. DOI: 10.1093/infdis/jix478.

32. Daulagala P, Leung YWY, Luk LLH, et al. Anti-neuraminidase antibody responses in older adults after consecutive vaccinations with enhanced influenza vaccines: a randomized controlled trial. J Infect Dis 2025. DOI: 10.1093/infdis/jiaf616.

33. Colombo RE, Richard SA, Schmidt K, et al. Randomized Pragmatic Trial of the Comparative Effectiveness of Chicken Egg-Based Inactivated, Mammalian Cell Culture-Based Inactivated, and Recombinant Protein Quadrivalent Seasonal Influenza Vaccines in United States Military Health System Beneficiaries. Clinical Infectious Diseases 2025;81(5):e454–e463. DOI: 10.1093/cid/ciaf503.

34. Lewis NM, Chung JR, Uyeki TM, Grohskopf L, Ferdinands JM, Patel MM. Interpretation of Relative Efficacy and Effectiveness for Influenza Vaccines. Clinical Infectious Diseases 2022;75(1):170–175. DOI: 10.1093/cid/ciab1016.

35. Krammer F. Next-generation seasonal influenza virus vaccines need a neuraminidase component. Vaccine 2025;54:126994. DOI: 10.1016/j.vaccine.2025.126994.

36. Monto AS, Petrie JG, Cross RT, et al. Antibody to Influenza Virus Neuraminidase: An Independent Correlate of Protection. Journal of Infectious Diseases 2015;212(8):1191–1199. DOI: 10.1093/infdis/jiv195.

37. Fitz-Patrick D, McVinnie DS, Jackson LA, et al. Efficacy, Immunogenicity, and Safety of Modified mRNA Influenza Vaccine. New England Journal of Medicine 2025;393(20):2001–2011. DOI: 10.1056/nejmoa2416779.

