## Supplemental Materials for "Comparative immunogenicity and relative vaccine efficacy of recombinant and cell culture-based influenza vaccines: Results from a randomized trial of adults in the United States, 2024-2025"

Bradley Bobbett,[3] Kelly Conard,[3] Raquel Salgado,[4] Adriana Rauseo Acevedo,[6] Stephanie Fritz,[6] Ryley Thompson,[6] Alem Haile,[6] Lisa Kessels,[6] Emily A. Randolph,[7] Darren J. Morrow,[7] Curtis Donskey, MD,[11] Joy Abou Farah, MD,[9] Alexia El Khoury,[9] Clarice Farley,[9] Jose Andres Carlos Portillo,[9] Todd M Bear,[12] Michael Susick,[12] M Patricia Nowalk,[12] Krissy Moehling Geffel,[12] John F Alcorn,[12] Peyton K. F. Sims,[12] Gregory Castelli[12]

### SUPPLEMENTAL METHODS

*Influenza A subtyping by serologic confirmation*

Influenza A subtyping was determined by RT-PCR testing or serologic confirmation. RT-PCR testing was used as the primary subtyping determination. Serologic confirmation was defined as a ≥4-fold rise in HAI or MN antibody titer or a ≥2-fold rise in binding antibody titer against a single influenza subtype from the acute illness blood draw (< 10 days after symptom onset) to the convalescent blood draw (28 days later). Serologic confirmation was concurrent with 90% (9/10) of those that had RT-PCR subtyping completed.

*Seroprotection and Seroconversion*

To evaluate additional immunogenicity outcomes for the trial's primary objective, we calculated the proportion of participants in each randomization group with a seroprotective HAI titer (≥1:40) post-vaccination as well as the proportion who achieved seroconversion by HAI titers following vaccination, defined as an HAI titer ≥1:40 if the baseline titer is <1:10 or a four-fold rise in HAI titer if the baseline titer is ≥1:10. A logistic regression model adjusted for study site compared seroconversion and seroprotection percentages across randomization group. For A(H3N2), MN titers were used in lieu of HAI titers and the same methods were applied.

*Serious Adverse Events (SAEs)*

An SAE was defined as an adverse event that meets one of the following conditions: results in death during the period of protocol-defined surveillance; is life-threatening (defined as immediate risk of death at the time of the event); requires inpatient hospitalization or prolonged hospitalization during the period of protocol-defined surveillance; results in congenital anomaly or birth defect; results in a persistent or significant disability/incapacity; or any other important medical event that may not result in death, be life threatening, or require hospitalization, may be considered an SAE when, based upon appropriate medical judgment, the event may jeopardize the participant and may require medical or surgical intervention to prevent one of the outcomes listed above. Examples of such medical events include allergic bronchospasm requiring intensive treatment in an emergency room or at home, blood dyscrasias or convulsions that do not result in inpatient hospitalization, or the development of drug dependency or drug abuse.[1]

SAEs resulting in hospitalization were identified via the weekly active surveillance survey where participants reported a change in health status requiring hospitalization in the past 7 days. Study staff followed up with participants that reported “yes” for this question for additional details to determine relationship to study vaccine or procedures. Participants may have also passively reported SAEs to study staff via email or phone.

### Supplemental References

1. US Food and Drug Administration. What is a Serious Adverse Event? (https://www.fda.gov/safety/reporting-serious-problems-fda/what-serious-adverse-event).

### SUPPLEMENTAL TABLES

**Supplemental table 1. Demographics of study participants in the immunogenicity subset**

| Characteristic | Randomization group | |
|---|---|---|
| | RIV | ccIIV |
| **Total N** | 76 | 75 |
| **Age (years), median (IQR)** | 46 (30, 55) | 37 (28, 45) |
| **Age Group, n (%)** | | |
| 18-49 years | 47 (61.8%) | 60 (80.0%) |
| 50-64 years | 29 (38.2%) | 15 (20.0%) |
| **Sex,[a] n (%)** | | |
| Female | 54 (71.1%) | 54 (72.0%) |
| Male | 20 (26.3%) | 21 (28.0%) |
| **Race/ethnicity, n (%)** | | |
| Hispanic or Latino | 10 (13.2%) | 9 (12.0%) |
| American Indian or Alaska Native | 0 (0.0%) | 2 (2.7%) |
| Asian | 3 (3.9%) | 11 (14.7%) |
| Black or African American | 6 (7.9%) | 8 (10.7%) |
| Middle Eastern or North African | 0 (0.0%) | 0 (0.0%) |
| Multiracial | 2 (2.6%) | 0 (0.0%) |
| Native Hawaiian or Other Pacific Islander | 0 (0.0%) | 0 (0.0%) |
| White | 53 (69.7%) | 45 (60.0%) |
| Other | 0 (0.0%) | 0 (0.0%) |
| Unknown | 2 (2.6%) | 0 (0.0%) |
| **Enrollment Site, n (%)** | | |
| Arizona State University | 17 (22.4%) | 17 (22.7%) |
| Valleywise Health Medical Center | 6 (7.9%) | 5 (6.7%) |
| Washington University | 19 (25.0%) | 19 (25.3%) |
| University Hospitals | 17 (22.4%) | 17 (22.7%) |
| Cleveland Veterans Affairs | 4 (5.3%) | 4 (5.3%) |
| University of Pittsburgh | 13 (17.1%) | 13 (17.3%) |
| **Prior season influenza vaccination, n (%)** | | |
| No | 5 (6.6%) | 8 (10.7%) |
| Yes | 70 (92.1%) | 66 (88.0%) |
| I don't know | 1 (1.3%) | 1 (1.3%) |
| **Current season influenza A infection,[b] n (%)** | | |
| Negative | 74 (97.4%) | 67 (89.3%) |
| Positive[c] | 2 (2.6%) | 8 (10.7%) |

Abbreviations: ccIIV, cell culture-based inactivated influenza vaccine; RIV, recombinant influenza vaccine
[a] Sex was unknown for 3 participants; 3 RIV
[b] Defined as a test-positive influenza A infection as indicated by a positive molecular or rapid diagnostic test result.
[c] Influenza confirmed by at-home antigen test for 2 participants; 1 RIV and 1 ccIIV

**Supplemental table 2. Primary objective comparing percent seroprotection and seroconversion conferred by RIV and ccIIV influenza vaccines in immunogenicity subset**

| Influenza antigen | RIV[a] | ccIIV[b] | *p*-value[e] comparing ccIIV to RIV |
|---|---|---|---|
| | Percent (95% CI) | Percent (95% CI) | |
| **Total n (row %)** | 76 (50.3%) | 75 (49.7%) | - |
| **A(H1N1)pdm09** | | | |
| Percent seroprotection[c] | 88.2 (78.6, 93.8) | 81.3 (70.7, 88.7) | 0.224 |
| Percent seroconversion[d] | 68.4 (57.0, 78.0) | 30.7 (21.2, 42.1) | <0.001 |
| **A(H3N2)** | | | |
| Percent seroprotection[c] | 94.7 (86.6, 98.0) | 82.7 (72.2, 89.7) | 0.022 |
| Percent seroconversion[d] | 86.8 (77.1, 92.8) | 45.3 (34.3, 56.8) | <0.001 |
| **B/Victoria** | | | |
| Percent seroprotection[c] | 86.8 (77.1, 92.8) | 74.7 (63.5, 83.3) | 0.057 |
| Percent seroconversion[d] | 48.7 (37.5, 60.0) | 32.0 (22.3, 43.5) | 0.035 |

Abbreviations: ccIIV, cell culture-based inactivated influenza vaccine; CI, confidence interval; RIV, recombinant influenza vaccine

Note: Viruses tested were cell-grown H1N1 A/WI/67/22 and BVic B/AUS/1359417/21 using hemagglutination inhibition and H3N2 A/MA/18/22 using microneutralization.

[a] RIV contains 45 µg of HA per strain (i.e. a total of 135 µg HA) and no NA

[b] ccIIV contains 15 µg of HA per strain, i.e. a total of 45 µg HA and varying NA content per strain; the lot given to participants was quantified as containing <0.5 µg for A(H1N1)pdm09, 10.4 µg for A(H3N2), and 1.5 µg for influenza B

[c] Seroprotection was defined as a post-vaccination hemagglutination inhibition or microneutralization titer (≥1:40).

[d] Seroconversion was defined as a post-vaccination hemagglutination inhibition or microneutralization titer ≥1:40 if the pre-vaccination titer is <1:10 or a four-fold rise in post-vaccination titer if the pre-vaccination titer is ≥1:10.

[e] Estimated using a logistic regression adjusted for site.

**Supplemental table 3. Sensitivity analyses of relative vaccine efficacy against influenza A overall, influenza A(H1N1)pdm09, and influenza A(H3N2) comparing receipt of RIV to ccIIV.**

| Sensitivity analysis | Endpoint | Influenza cummulative incidence | | Relative vaccine efficacy |
|---|---|---|---|---|
| | | RIV[a] | ccIIV[b] | |
| | | % | | % (95% CI) |
| Main analysis against test-positive influenza illness[c] | **Overall influenza A** | 2.4 | 6.1 | 61 (7 to 84) |
| | **Influenza A subtype** | | | |
| | A(H1N1)pdm09 | 0.3 | 2.7 | 87 (-1 to 98) |
| | A(H3N2) | 1.0 | 3.0 | 66 (-24 to 91) |
| Sensitivity analysis against test-positive influenza illness with allowing for stratified baseline hazard by enrollment site[d] | **Overall influenza A** | 2.4 | 6.1 | 61 (7 to 84) |
| | **Influenza A subtype** | | | |
| | A(H1N1pdm09) | 0.3 | 2.7 | 87 (-1 to 98) |
| | A(H3N2) | 1.0 | 3.0 | 67 (-24 to 91) |
| Sensitivity analysis against self-reported influenza illness[e] | **Overall influenza A** | 3.7 | 7.7 | 52 (2 - 77) |
| Sensitivity analysis againgst RT-PCR-positive influenza illness[f] | **Overall influenza A** | 2.0 | 6.5 | 69 (15 to 89) |
| | **Influenza A subtype** | | | |
| | A(H1N1)pdm09 | 0.4 | 2.4 | 83 (-39 to 98) |
| | A(H3N2) | 1.2 | 3.2 | 62 (-42 to 90) |

Abbreviations: ccIIV, cell culture-based inactivated influenza vaccine; CI, confidence interval; RIV, recombinant influenza vaccine; RT-PCR, reverse transcription-polymerase chain reaction

[a] RIV contains 45 µg of HA per strain (i.e. a total of 135 µg HA) and no NA

[b] ccIIV contains 15 µg of HA per strain, i.e. a total of 45 µg HA and varying NA content per strain; the lot given to participants was quantified as containing <0.5 µg for A(H1N1)pdm09, 10.4 µg for A(H3N2), and 1.5 µg for influenza B

[c] Relative vaccine efficacy of RIV compared to ccIIV from a Cox proportional hazards model of the time to test-positive influenza illness adjusted for influenza vaccine type. Analysis includes 591 participants.

[d] Relative vaccine efficacy of RIV compared to ccIIV from a Cox proportional hazards model of the time to test-positive influenza illness adjusted for influenza vaccine type with a strata term for enrollment site. Analysis includes 591 participants.

[e] Relative vaccine efficacy of RIV compared to ccIIV from a Cox proportional hazards model of the time to test-positive or self-reported influenza illness adjusted for influenza vaccine type. Analysis includes 591 participants.

[f] Relative vaccine efficacy of RIV compared to ccIIV from a Cox proportional hazards model of the time to RT-PCR-positive influenza illness adjusted for influenza vaccine type. One enrollment site (Pittsburgh, PA) did not do RT-PCR testing and was excluded for these analyses. Analysis includes 492 participants.

**Supplemental table 4. Participants reporting serious adverse events (SAE) during the study period.**

| Vaccination group | Age group | Time since vaccination (days) | Relatedness | SAE category | SAE description |
|---|---|---|---|---|---|
| ccIIV | 50-64 years | 153 | Not related | Death | Stage IV lung cancer |
| ccIIV | 18-49 years | 170 | Not related | Hospitalization | Kidney stones |
| ccIIV | 18-49 years | 199 | Not related | Important medical event | Whip lash from motor vehicle accident |

Abbreviations: ccIIV, cell culture-based inactivated influenza vaccine
